# Prevalence of Mild Cognitive Impairment in the Lothian Birth Cohort 1936

**DOI:** 10.1101/2020.10.08.20209130

**Authors:** Miles Welstead, Michelle Luciano, Graciela Muniz-Terrera, Adele M. Taylor, Tom C. Russ

**Affiliations:** Lothian Birth Cohorts, Department of Psychology, University of Edinburgh, 7 George Square, Edinburgh, UK; Edinburgh Dementia Prevention, University of Edinburgh, BioCube 1, Edinburgh, UK; Alzheimer Scotland Dementia Research Centre, 7 George Square, University of Edinburgh, Edinburgh, UK

**Keywords:** MCI, Cognitive ageing, Amnestic, Non-amnestic, Prevalence

## Abstract

**Background:** The Lothian Birth Cohort 1936 (LBC1936) is a highly-phenotyped longitudinal study of cognitive and brain ageing. Given its substantial clinical importance, we derived an indicator of mild cognitive impairment (MCI) as well as amnestic and non-amnestic subtypes at three time points.

**Methods:** MCI status was derived at three waves of the LBC1936 at ages 76 (*n*=567), 79 (*n*=441), and 82 years (*n*=341). A general MCI category was derived as well as amnestic MCI (aMCI) and non-amnestic MCI (naMCI). A comparison was made between MCI derivations using normative data from the LBC1936 cohort versus the general UK population.

**Results:** MCI rates showed a proportional increase at each wave between 76 and 82 years from 15% to 18%. Rates of MCI subtypes also showed a proportional increase over time: aMCI 4% to 6%; naMCI 12% to 16%. Higher rates of MCI were found when using the LBC1936 normative data to derive MCI classification rather than UK-wide norms.

**Conclusions:** We found that MCI and aMCI rates in the LBC1936 were consistent with previous research. However, naMCI rates were higher than expected. Future LBC1936 research should assess the predictive factors associated with MCI prevalence to validate previous findings and identify novel risk factors.

## Background

In conjunction with advancements in health and social care in the past century, life expectancy has improved dramatically and contributed to a rapidly increasing older population.^1^ A consequence of this demographic shift is the challenge we now face to care for a larger number of older adults with susceptibility to cognitive deterioration.^2^ Understanding how cognitive decline affects older people is imperative in order to design interventions to slow or delay decline and ensure individuals are on the healthiest ageing trajectory possible.^3^ Decline in memory is a key indicator of dementia, however it is common in older age, and differences between normal age-related decline and the early stages of dementia can be difficult to differentiate.^4^

The concept of mild cognitive impairment (MCI) traces back many years but has gained particular traction over the past few decades.^5^ Petersen, Doody, Kurz, et al. ^6^ popularised the concept as a distinct clinical condition and established a set of criteria based on memory changes without loss of ability to undertake normal activities. These criteria heavily influenced the way in which MCI was, and continues to be, identified in research and clinical settings. However, other researchers such as Dubois & Albert ^7^ disputed the notion of MCI as a distinct clinical entity, instead proposing it as a stage of severity for particular disorders. Accordingly, they proposed a ‘prodromal Alzheimer’s Disease’ based upon subjective memory complaints with progressive onset, preserved ability to undertake activities of daily living, neuroimaging, and biomarker testing. Disagreement on how MCI should be conceptualised has led to multiple attempts at an international consensus. Winblad, Palmer, Kivipelto, et al. ^8^ reached consensus that MCI criteria should assess whether an individual has a dementia diagnosis, whether their cognition has shown subjective and/or objective decline over time, and whether their activities of daily living are significantly affected – and, indeed, how this latter criterion is judged. This groundwork informed the most recent guidelines proposed by the National Institute on Aging-Alzheimer’s Association (NIA-AA) workgroups on diagnostic guidelines for Alzheimer’s disease.^9^ These guidelines propose four criteria based on: 1. Concern regarding a change in cognition, 2. Impairment in one or more cognitive domains, 3. Preservation of independence in functional abilities, 4. No diagnosis of dementia. In addition to identifying general MCI, there has also been increased interest in identifying specific subtypes of MCI that may precede certain types of dementia. For instance, amnestic MCI (aMCI) focuses solely on memory-related cognitive impairment, whereas non-amnestic MCI (naMCI) focusses on cognitive impairment in other domains such as processing speed, attention, and executive functions.^10^ Whilst aMCI is associated with a high risk of converting to Alzheimer’s disease, naMCI is associated with other types of dementia such as diffuse Lewy body dementia.^11^ Identifying MCI in general as well as its subtypes will allow for improved knowledge on how early prevention strategies can identify individuals who are at high risk of cognitive decline and subsequent dementia. Here we use the NIA-AA guidelines to derive an identification of MCI and its subtypes using data from the Lothian Birth Cohort 1936.^12,13^ We hypothesise that MCI rates will be similar to those found in other older adult cohorts and that prevalence of all types of MCI will be higher in later data waves.

## Methods

At Wave 1, the LBC1936 study consisted of 1091 participants, born in 1936 with a mean age of 69 (SD=0.89) years, mostly surviving members of the Scottish Mental Survey 1947.^14^ Wave 1 took place between 2004 and 2007, with follow-up waves approximately every three years thereafter at ages: 73 (*n*=866), 76 (*n*=697), 79 (*n*=550), and 82 years (*n*=431). More details on recruitment and testing procedures have been published previously.^12,13,15^ The LBC1936 study was conducted according to the Declaration of Helsinki guidelines. Ethical permission for the LBC1936 study protocol was obtained from the Multi-Centre Research Ethics Committee for Scotland (Wave 1: MREC/01/0/56), the Lothian Research Ethics Committee (Wave 1: LREC/2003/2/29), and the Scotland A Research Ethics Committee (Waves 2, 3, 4 & 5: 07/MRE00/58). Written consent was obtained from participants at each of the waves.

### Identification of MCI

Using data previously collected in the LBC1936, an algorithm was created which identifies participants who fulfil the MCI criteria as outlined by the NIA-AA workgroups on diagnostic guidelines for Alzheimer’s disease.^9^ Variables necessary to conduct MCI coding were collected from Wave 3 (age 76) onwards. In order to be classified in the MCI category, participants must have shown met all four criteria reported below:

1. **Concern regarding a change in cognition:** Self-reported memory problems that are interfering with their life, as recorded in a questionnaire at each wave.
2. **Impairment in one or more cognitive domains:** Scores at least 1.5 SD below the mean on at least one cognitive domain (memory, executive function, attention, language, or visuospatial skills) ***AND*** either shows a decline from the previous wave to below the 10th percentile on one test, a decline from wave 1 to below the 20^th^ percentile on one test, or a decline from the previous wave to below the 20th percentile on two tests.
3. **Preservation of independence in functional abilities:** Scores at least 1.5 SD below the mean on the Townsend’s Disability Scale overall score.^16^
4. **No diagnosis of dementia:** Does not self-report or have a formal diagnosis of dementia ***AND*** scores at least 24 on the Mini-Mental State Examination (MMSE).^17^

### Cognitive domains were assessed using the following cognitive tests

Symbol Search, Digit Symbol Coding, Matrix Reasoning, Letter-Number Sequencing, and Block Design from the Wechsler Adult Intelligence Scale III (WAIS) and Logical Memory I & II from the Wechsler Memory Scale III (WMS-III).^18^ A cut-off of ≥1.5 SD below the mean or scoring below specific percentiles was used to indicate cognitive impairment. Two versions of the cognitive impairment criterion were conducted using the means and standard deviations of individual tests from (1) the LBC1936 sample at each wave and (2) a more representative UK sample provided by the WAIS-III-WMS-III technical manual.^18^ Preliminary comparisons showed that fewer participants were identified as having MCI using the general population norms, likely due to the higher rates of overall healthiness in the LBC1936.^13^ Therefore, the definition using UK normative data were used here as they were more reflective of the general population.

### We also coded two subtypes of MCI

Amnestic MCI (aMCI) and Non-amnestic MCI (naMCI). Creation of these subtypes followed the same procedure as for the general MCI, however aMCI was only identified if the participant showed impairment in the memory domain. Similarly, classification for naMCI was met if the participant showed impairment in cognitive domains other than memory (executive function, attention, language, or visuospatial skills).

### Covariates

We examined the association between a range of covariates and MCI status. Covariates included: age, sex, years of education, age 11 cognitive function, body mass index (BMI; calculated in the standard way of kg/m^2^), occupational social class (professional/managerial/skilled, non-manual/skilled manual or semiskilled/unskilled), *APOE* ε4 status (allele present/absent), self-reported history of cardiovascular disease, self-reported history of stroke, depression, and physical frailty level (not frail/pre-frail/frail). Physical frailty was derived using the Fried Phenotype guidelines^19^, for information on how this was calculated in LBC1936 see Welstead, Muniz-Terrera, Russ, et al. ^20^. Depression was measured using the Hospital Anxiety and Depression scale (HADS).^21^ Age 11 cognitive function was based on LBC1936 participant’s scores on the Moray House Test (MHT) at age 11^22^; for more detail see Taylor, Pattie, Deary ^13^. To adjust for age in days at time of testing, MHT11 scores were residualised for age at 11 years.

### Statistical Analysis

Three participants had been diagnosed with dementia before age 76 (wave 3) by the LBC1936 study doctor and were excluded, leaving 694 participants at that wave. Additionally, since a wide variety of variables were required in order to derive an MCI coding, missing data at each wave meant that some participants were excluded from analyses (wave 3; *n*=127, wave 4; *n*=106, wave 5; *n*=87). Accordingly, MCI status was coded for 567 participants at wave 3 (age 76), 441 at wave 4 (age 79), and 341 at wave 5 (age 82). Descriptive analyses including number and percentages of people with MCI were used to characterise the study sample. Linear Model ANOVAs and Pearson’s Chi-squared tests were used to assess characteristics associated with MCI and Non-MCI participants. All statistical analyses were conducted in R Version 3.6.1.^23^

## Results

**Figure 1** show the rates of MCI in the LBC1936. There was an increase in people with MCI over time with 15% at wave 3 (*n*=87/567), 17% at wave 4 (*n*=77/441), and 18% at wave 5 (*n*=62/341) having MCI. As there were a substantial number of participants who withdrew from the study between baseline and final follow-up, we also looked at MCI rates for completers only, i.e. those who completed waves 3, 4, and 5. Results showed an overall proportional increase over follow-up with 14% of completers identified as having MCI at wave 3 (n=38/271) and wave 4 (n=38/271), and then a rise to 21% at wave 5 (n=57/271).

**Figure 1:**
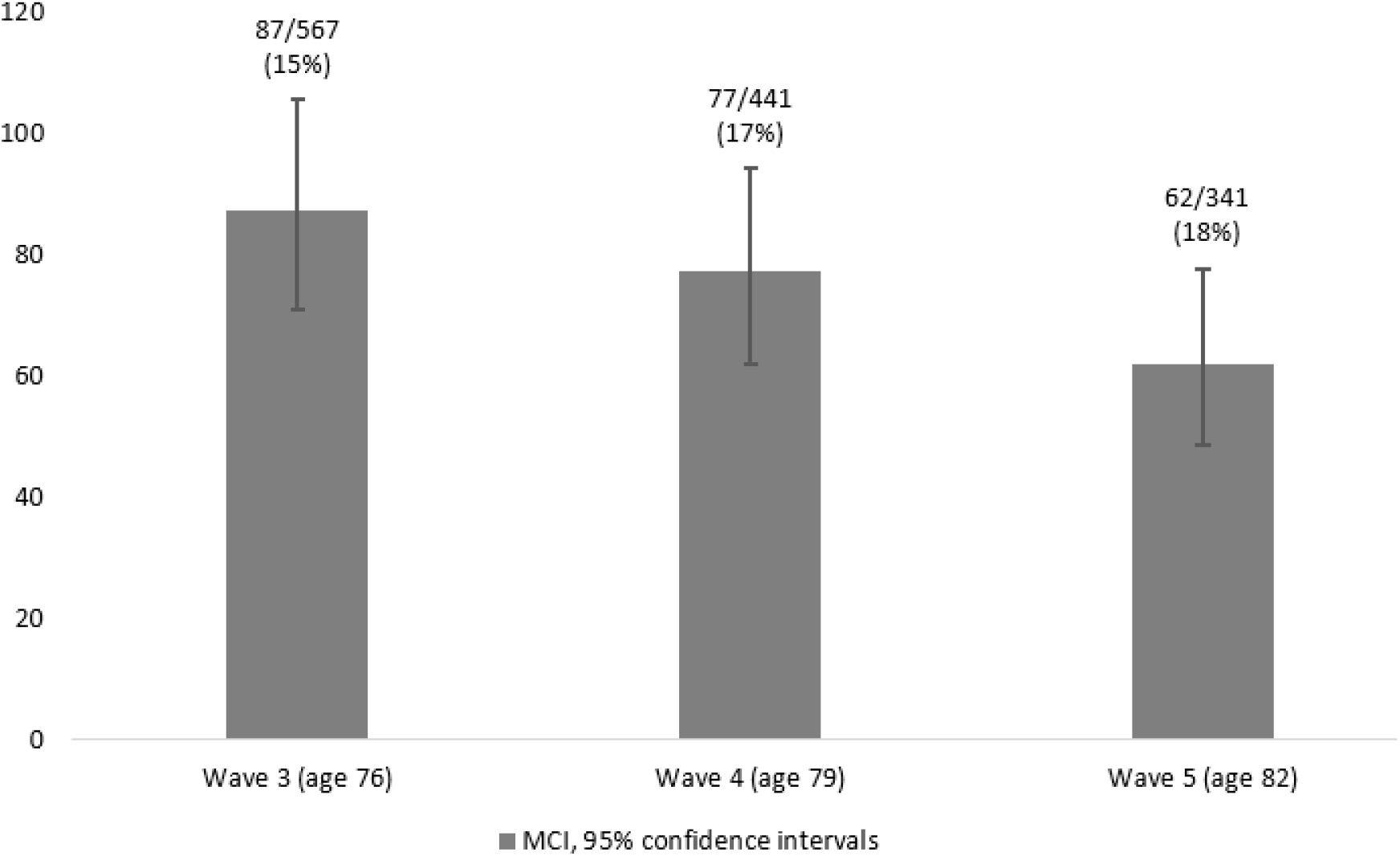
Comparisons of MCI rates in the Lothian Birth Cohort 1936 study across waves using UK wide normative data

MCI rates did not differ significantly by sex at any of the waves. The only significant differences found indicated that higher rates of MCI were associated with *APOE* ε4 status at wave 3 (*p*<0.001) and wave 5 (*p*<0.05), and history of stroke at wave 3 (*p*<0.01) and wave 5 (*p*<0.05). Covariate differences according to MCI status are reported in **Table 1**.

**Table 1:**
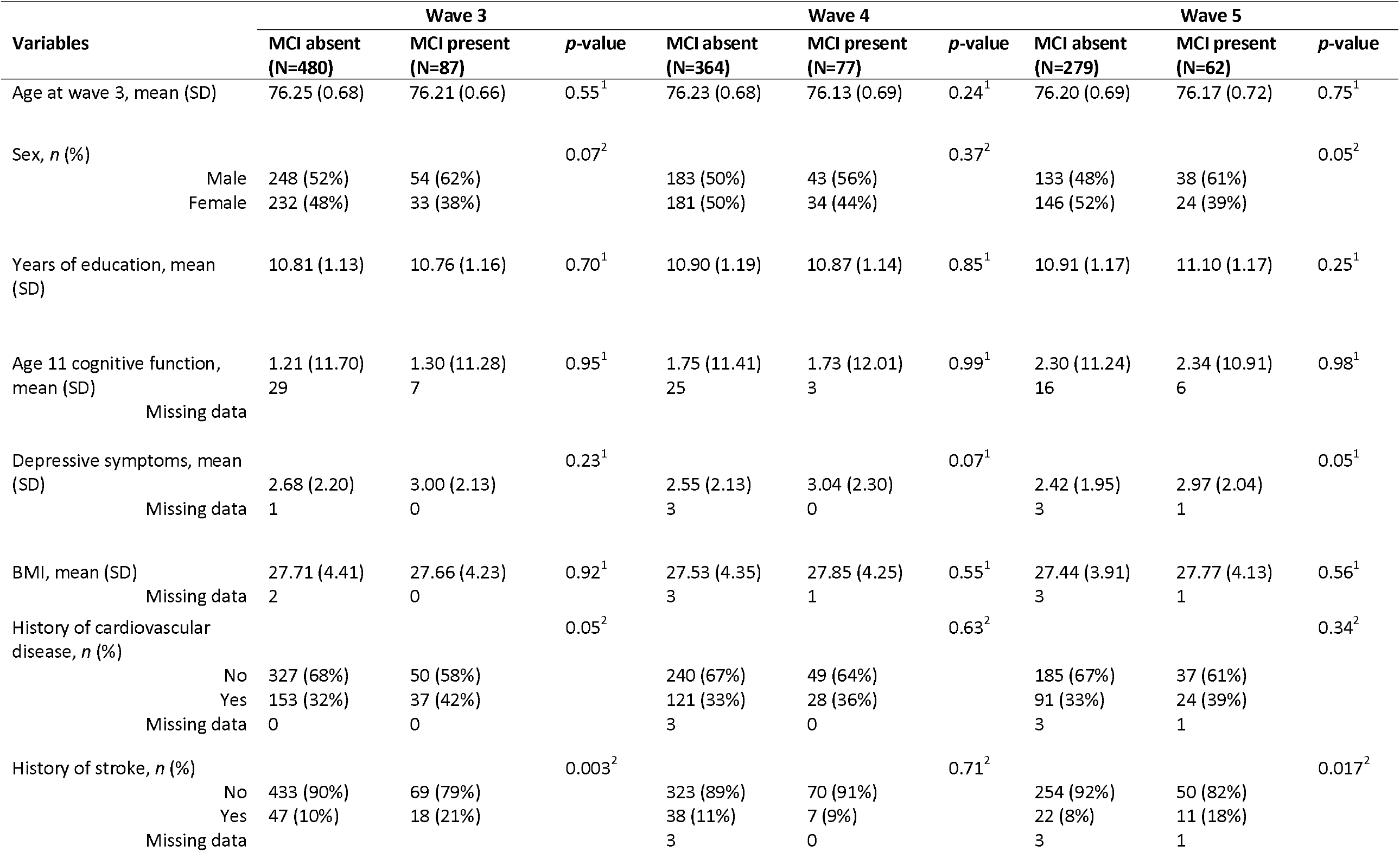
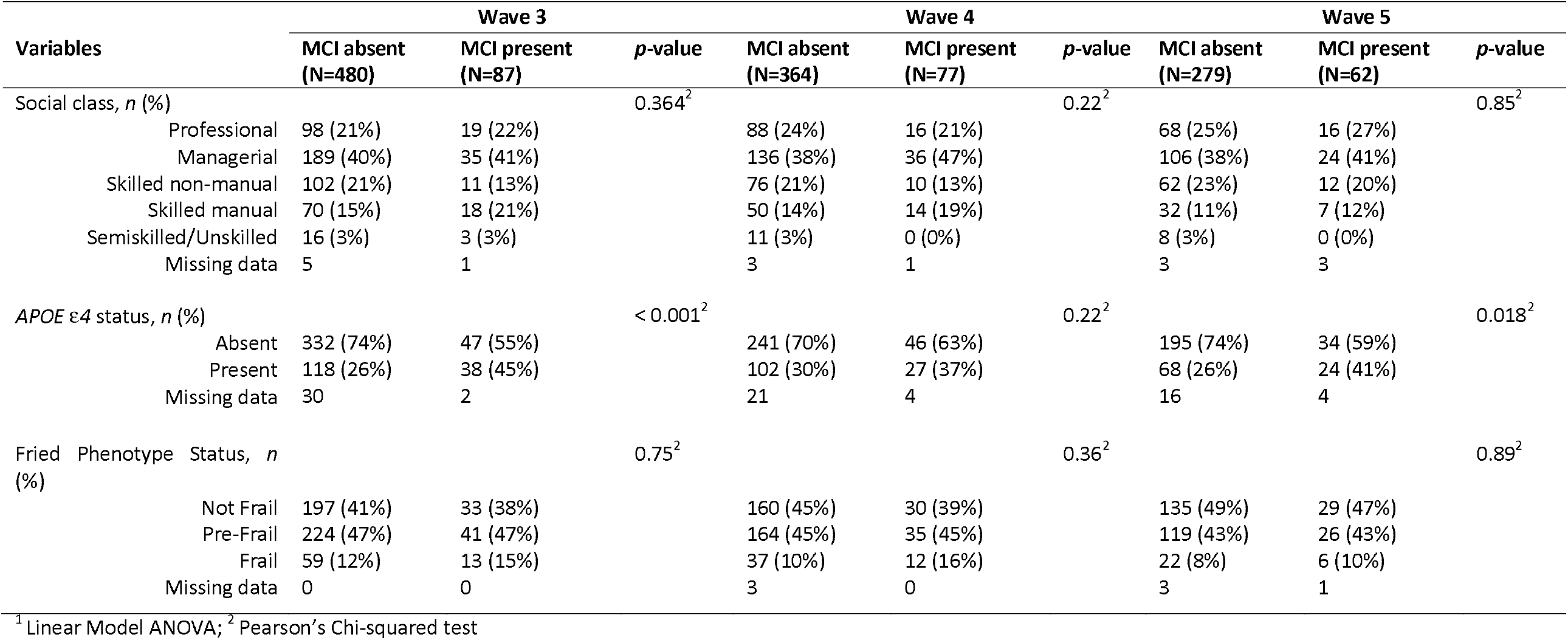
Covariate descriptive statistics for participants with MCI present vs absent.

### MCI subtypes

We also derived two subtypes of MCI: aMCI and naMCI. As reported in **Figure 2**, proportions of aMCI remained fairly low across follow-up from 4% at wave 3 (n=24/604), to 4% at wave 4 (n=21/484), and 6% at wave 5 (n=24/376). Prevalence of naMCI was higher and showed a gradual proportional increase over follow-up from 12% at wave 3 (n=73/609), to 14% at wave 4 (n=63/466), and 16% at wave 5 (n=56/361).

**Figure 2:**
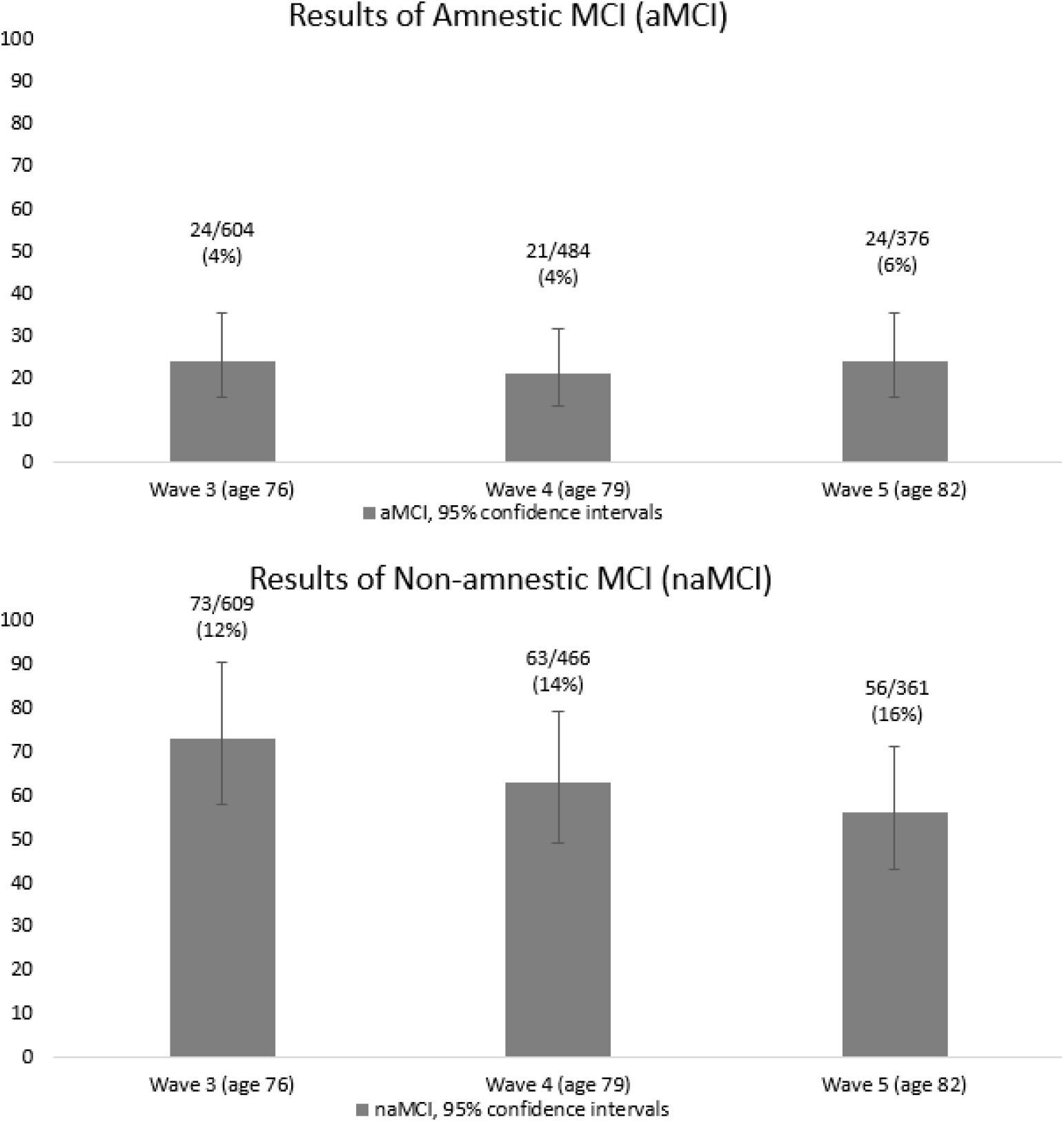
Comparisons of aMCI vs Non-aMCI rates in the Lothian Birth Cohort 1936 study across waves

### Normative data comparisons

We compared whether MCI rates were sensitive to the use of different normative data. Comparisons were made between MCI rates when using normative data based on the LBC1936 and a UK-wide sample to derive the identification of MCI. As might be expected with a healthy cohort, at all waves there were higher proportions of MCI when using the LBC1936 norms compared to the UK based norms. **Supplementary figure 1** reports MCI rates at each wave according to the LBC1936 normative data.

## Discussion

We found MCI proportions in the LBC1936 study of 15%, 17%, and 18% at ages 76, 79, 82 years, respectively. Similar proportions were found when looking only at the individuals who attended all waves. MCI status at wave 3 and wave 5 (but not wave 4) was significantly associated with *APOE* ε4 status and history of stroke. Proportions of people with aMCI were 4% at ages 76 and 79 years and 6% at 82 years, whereas rates of naMCI were higher but still showed an increase in proportions from 12% at age 76 years to 14% and 16% at 79 and 82 years, respectively.

### Comparison with other literature

We observed higher rates of MCI in men, albeit not at a statistically significant level, a finding that is consistent with some previous research^24,25^, but not all.^26,27^ As discussed by Xue, Li, Liang, Chen ^27^, sex differences in MCI research are inconsistent and may differ according to alternate methods of deriving MCI. Importantly, the assessment of day-to-day function in men and women presents different challenges, and perhaps surprisingly, there were minimal significant associations between groups of individuals defined by key features. At two of the time points *APOE* ε4 status was associated with having MCI, a finding which has been consistently found in previous MCI research and is also strongly linked to the risk of progression to dementia.^28^ The only other characteristic associated with MCI change was having a history of stroke, again somewhat unsurprising given the extensive evidence that stroke patients have higher risk for developing of MCI and dementia.^29^ The lack of significant association between these factors and MCI status at wave 4 is unexpected and not readily explained. However, it may be related to attrition or other factors leading to sample differences at wave 4; the proportion of participants with MCI who had an *APOE* ε4 allele present or a history of stroke was lower at wave 4 than waves 3 or 5.

As expected, findings also showed an increase in proportion of participants with MCI at wave 5 compared to wave 3. The rates of MCI we find are consistent with previous research using the same MCI coding guidelines which reports an average prevalence of 14.8% for 70-75 year olds.^30^ The rates of two subtypes of MCI – aMCI and naMCI – were in partial agreement with previous literature. Some previous research^10^ has found rates of around 3-4% of both aMCI and naMCI in older populations, whilst others have found 11% for aMCI and 5% prevalence for naMCI.^26^ Thus, whilst the aMCI results are expected, the rates of naMCI in the LBC1936 are higher than anticipated. Higher rates of naMCI than aMCI may indicate that participants of the LBC1936 are more prone to non-amnestic cognitive impairment in areas such as language, visual-spatial skills, attention, or executive functioning. Another possibility is that the salient memory problems associated with aMCI may make participants more likely to withdraw from the study, whereas the cognitive problems associated with naMCI (executive function, attention, language, or visuospatial skills) may more often go unnoticed by the participant. However, it is also important to note that making comparisons between our proportions of aMCI and naMCI cannot be done entirely accurately given that cases of missing data differed between them.

### Limitations and Strengths

LBC1936’s rates of high physical health and cognitive ability is well documented^13,15^, and highlights a limitation of this study: our sample is less representative of the general population who likely have higher rates of MCI. An additional limitation that affects the accuracy of our results was that there was a relatively small number of participants who were identified as having aMCI, which introduces an element of uncertainty into our results. For the participants who withdrew from the study, we did not have systematic information on their reason for dropping out. It is likely that at least some of these participants dropped out due to MCI or dementia, and accordingly we were unable to consider these cases in our analyses. Related to this, other than three cases in which we had confirmation from the LBC1936 study doctor, we relied primarily on the self-reporting of dementia diagnoses for part of the MCI criteria. This could have introduced bias if additional participants had a dementia diagnosis but did not report it. Whilst self-reporting is used extensively in epidemiological studies and biases are usually insignificant^31^, given the nature of dementia, using these measures may have introduced inaccuracies. Current work is being undertaken in the LBC1936 to ascertain dementia status for every participant and so future research will be able to revisit this.

The strengths of this study are our use of data collected at multiple time points over the course of approximately six years in a well-characterised longitudinal cohort study. Using more than one time point gives us better insight into how MCI proportions change over time in the LBC1936. An additional strength is that we derived and compared an MCI coding using normative cognitive data from the LBC1936 sample and the UK wide norms. By doing so, we were able to assess the extent to which the LBC1936 data are representative of the wider population. As anticipated, MCI rates were higher at all waves when using the LBC1936 norms, presumably due to an overestimation caused by the higher rates of healthiness found in the LBC1936 when compared to the general population. Deriving MCI using the cohort’s own normative data will cause the cognitive impairment cut-off points to be more lenient than using normative data from the UK population as we see in our results.

### Implications

Our findings have added to the current literature by providing information on the prevalence of MCI in a prominent longitudinal cohort study, reinforcing findings found in similar cohorts. The identification of individuals with MCI in the LBC1936 and their comparison with findings in similar cohorts provides opportunities for future research to further explore MCI in this cohort. In particular, utilising the wealth of longitudinal data in the LBC1936 could prove insightful. MCI has been shown to be relatively fluid over time with both declines and reversions being common^32-34^. Accordingly, understanding this fluidity and the predictive factors associated with MCI change will be insightful for future interventions and prevention strategies that aim to lower the risk of MCI developing and progressing.

## Conclusion

This study is largely consistent with previous research, finding MCI rates of 15% to 18% in the LBC1936 at ages 76 to 82. When considering subtypes of MCI, non-amnestic MCI is more likely to affect participants than amnestic MCI indicating that perhaps this population is more prone to cognitive decline in non-amnestic cognitive domains. These results help highlight the prevalence of MCI in the LBC1936 and allow for future studies to explore cognitive trajectories over time and the predictive factors which may increase the risk of developing MCI.

## Data Availability

R script can be provided upon request

