## Supplementary figure 1 for "Prevalence of Mild Cognitive Impairment in the Lothian Birth Cohort 1936"

Supplementary figure 1: MCI rates using normative data from the Lothian Birth Cohort 1936

|  | MCI, 95% confidence intervals |
| --- | --- |
| Wave 3 (age 76) | 104 |
| Wave 4 (age 79) | 96 |
| Wave 5 (age 82) | 73 |
